# Genetic determinants of trehalose utilization are not associated with severe *Clostridium difficile* infection

**DOI:** 10.1101/19008342

**Authors:** Katie Saund, Krishna Rao, Vincent B. Young, Evan S. Snitkin

## Abstract

In a case-control study of patients with *C. difficile* infection we found no statistically significant association between the presence of trehalose utilization variants in infecting *C. difficile* strains and development of severe infection. These results do not support trehalose utilization conferring enhanced virulence in the context of human *C. difficile* infections.

## INTRODUCTION

*Clostridium difficile* infection (CDI) is a health care-associated infection that can result in a range of patient outcomes. Of greatest concern is the development of severe disease, defined as intensive care unit admission, intraabdominal surgery (such as colectomy), and/or death attributable to the infection[1]. Specific patient factors such as age, antibiotic use, and female gender have been associated with severe infection. The genetic background of the infecting *C. difficile* isolate may also influence clinical outcome. Prior studies reported an increased risk of severe infection for ribotypes RT027 and RT078[2,3]. One recently proposed mechanism for increased virulence of specific *C. difficile* lineages is the differential capacity to utilize the dietary disaccharide trehalose[4]. These authors observed enhanced trehalose utilization in strains of both RT027 and RT078, and demonstrated increased virulence of an RT027 strain in a mouse model of infection when physiologically relevant quantities of trehalose were given. This report also noted the coincidence between the introduction of trehalose as a food additive in 2000 and the global emergence of RT027 and RT078 shortly thereafter.

Subsequently, Eyre et al. examined clinical *C. difficile* isolates for the ability to use trehalose, noting that variants conferring improved trehalose utilization were not confined to successful epidemic lineages. Moreover, Eyre et al. found no evidence of association between a trehalose utilization variant and 30-day all-cause mortality in RT015, a ribotype in which enhanced trehalose utilization is variably present[5]. Here, we set out to more comprehensively evaluate the potential contribution of trehalose utilization to clinical outcomes by quantifying the independent contribution of trehalose utilization variants to infection severity across ribotypes, when controlling for all clinical factors independently associated with risk for severe infection.

## METHODS

### Study population

All subjects signed written consent to participate in this study. This study was approved by the University of Michigan Institutional Review Board (Study HUM33286). We considered a cohort of 1144 cases of CDI from hospitalized adults diagnosed with CDI between October 2010 and January 2013 at the University of Michigan Health System[2]. The following predictors of severe outcome were noted in the window 24-48 hours post CDI diagnosis: age, gender, metastatic cancer, concurrent antibiotic use, systolic blood pressure, creatinine, bilirubin, and white blood cell count. Of the 981 unique patients, 898 had complete clinical information. CDI was classified as severe if any of the following outcomes attributable to CDI occurred within 30 days of diagnosis: admission to an intensive care unit, intraabdominal surgery, or death[1]. Ribotyping was performed and 137 ribotypes were identified (Supplementary Table 1).

### Data analysis

Summary statistics, matching, modeling, and imputation were conducted in R version 3.5.0[6]. The R code is available at: github.com/katiesaund/clinical_cdifficile_trehalose_variants.

#### Severe Outcome Risk Score Matchin

Unique subjects with complete clinical information (N=898) were assigned a severe outcome risk score based on a logistic regression model with severe CDI outcome as the response variable and the following predictors: age (years), female gender, metastatic cancer, concurrent antibiotic use, systolic blood pressure (mm Hg), creatinine (>1.5 mg/dL), bilirubin (>1.2 mg/dL), and white blood cell count (cells/µL)[7]. Where possible, isolates were sorted into strata, each with exactly one case (severe CDI outcome) and at least one and up to four controls (non-severe CDI outcome), with control scores within ± 0.10 of case score. The matching process identified N=323 CDI cases isolates, of which only N=247 C. difficile isolates were successfully grown, isolated, and sequenced. Due to growth or sequencing failures only N=235 isolates were from N=59 complete strata (strata with at least one control and one case).

#### Conditional logistic regression model for matched samples

Logistic regressions were modeled with severe CDI outcome as the response variable, conditioned on strata, and a trehalose variant as the predictor. Bonferroni corrected P-values were reported.

### Genomic Analysis

Genomic DNA extracted with the MoBio PowerMag Microbial DNA Isolation Kit (Qiagen) from *C. difficile* isolates (N=247) was prepared for sequencing using the Illumina Nextera DNA Flex Library Preparation Kit. Sequencing was performed on either an Illumina HiSeq 4000 System at the University of Michigan Advanced Genomics Core or on an Illumina MiSeq System at the University of Michigan Microbial Systems Molecular Biology Laboratories. Quality of reads was assessed with FastQC[8]. Adapter sequences and low-quality bases were removed with Trimmomatic[9]. Details on sequenced strains are available in Table S1. Sequence data are available under Bioproject PRJNA561087. For variant calling details see the Supplementary Methods. Pangenome analysis was performed with roary[10] and the insertion putatively conferring enhanced trehalose utilization was inferred based on the presence of four genes: *treA2, ptsT, treX*, and *treR2* (Supplementary File 1).

## RESULTS

Given the limited clinical data regarding the potential contribution of trehalose utilization variants to CDI severity, we set out to test for an association between trehalose utilization across all strains of *C. difficile* causing infection while comprehensively controlling for patient factors associated with severe outcome. Patient factors associated with CDI severity in our clinical cohort (N=1144) were age, female gender, metastatic cancer, concurrent antibiotic use, systolic blood pressure, creatinine, bilirubin, and white blood cell count[2]. To quantify the independent contribution of trehalose utilization variants to severe patient outcomes we implemented a retrospective, matched case-control study to control for these patient factors. Each *C. difficile* episode was assigned a severe outcome risk score utilizing variables available around the time of diagnosis, which is the patient’s predicted probability of having severe CDI, based on a logistic regression model of CDI built from the eight patient factors. Unique patients were grouped into strata based on their risk score. Each stratum contained one case (N=59, severe CDI outcome) and up to four controls (N=176, non-severe CDI outcome; total N=235). The median number of controls per case was 3 (range, 1-4). There were 61 RT027 isolates (19/59 cases; 46/176 controls), 5 RT078 isolates (0/59 cases; 5/176 controls), and only 1 RT015 (1/59 cases; 0/176 controls) in this matched data set.

All of the *C. difficile* isolates in the matched data set with complete strata were sequenced (N=235). We identified the presence of reported trehalose utilization variants: two amino acid substitutions in the transcriptional repressor, *treR*, (TreR C171S and TreR L172I) and a set of four accessory genes, called the four gene trehalose insertion, that contain an additional phosphotrehalase enzyme and transcriptional regulator[4,11]. Consistent with previous reports, we found TreR C171S in all 4 RT017 isolates and 3 closely related ribotypes (Figure 1). Similarly, we found TreR L172I in all 61 RT027 isolates and in 7 closely related ribotypes. However, in contrast to Collins et al. who found the four gene trehalose insertion only in RT078, we identified the insertion in 25 isolates that were broadly distributed across the phylogeny[4].

**Figure 1.**
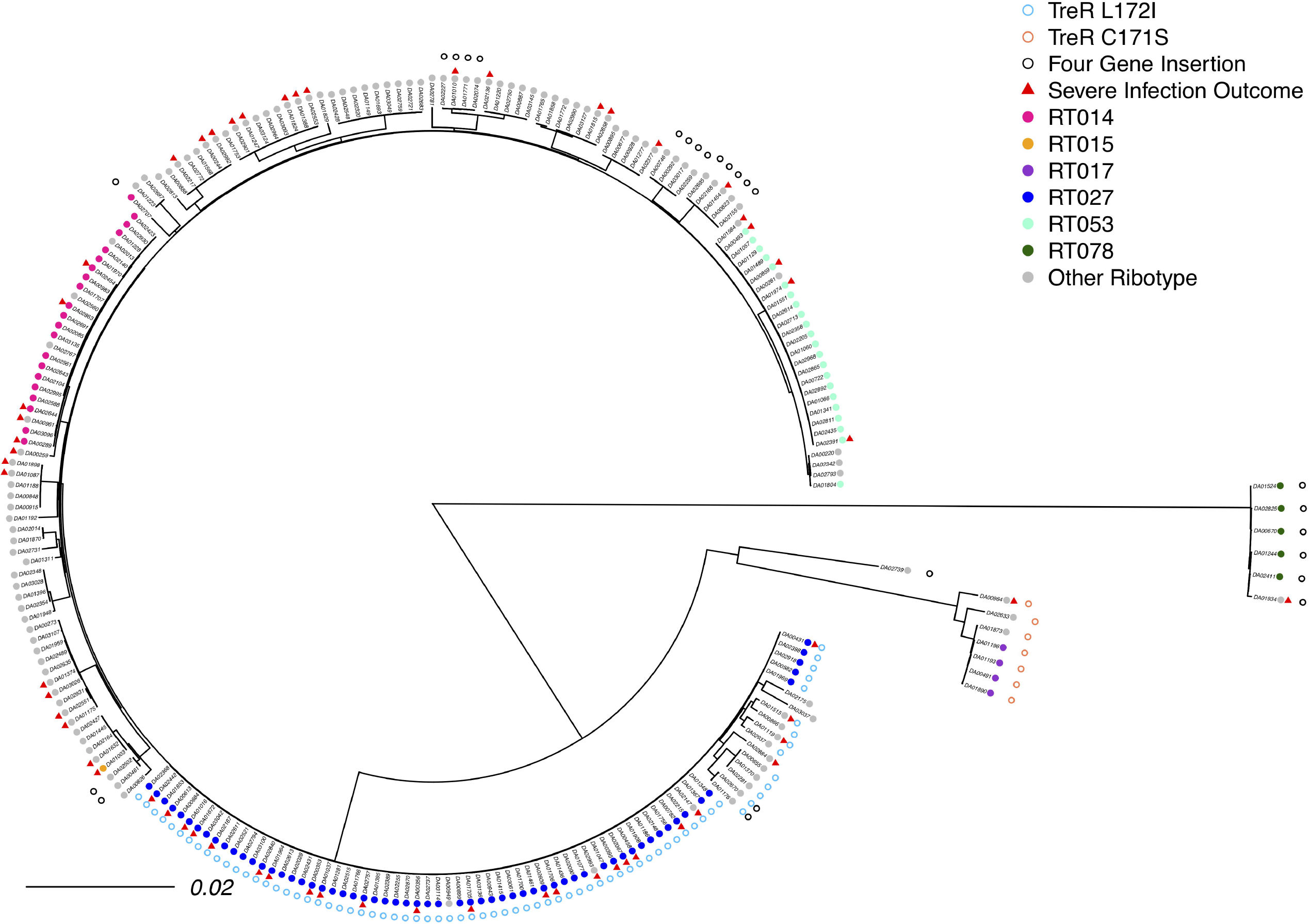
Comparative analysis of trehalose genetic variants in *Clostridium difficile*. A maximum-likelihood phylogeny was constructed (N=235) using variants identified in the genome (scale in mutations per site in whole genome). Trehalose variants are either point mutations in *treR* (C171S empty orange circle, L172I empty blue circle) or presence of the four gene trehalose insertion (empty black circle). A severe infection outcome is indicated by a red triangle. Ribotype is indicated by the filled circles.

To investigate if trehalose utilization variants were individually associated with severe CDI outcome, we performed unadjusted logistic regressions conditioned on severe outcome risk strata (Table S2). The presence of the TreR L172I trehalose utilization variant was associated with an increased, but not statistically significant, odds of severe CDI outcome (OR, 1.60; 95% CI, 0.84-3.05; *P*=0.52). The presence of the four gene trehalose insertion was associated with decreased, but not statistically significant odds of a severe CDI outcome (OR, 0.35; 95% CI, 0.09-1.32; *P*=0.52). None of the other four amino acid substitutions observed in TreR were significantly associated with severe CDI outcome (Table S2). Due to low prevalence (N=7), we did not test for an association between the presence of the TreR C171S trehalose utilization variant and severe CDI outcome.

To evaluate the overall effect of the three previously described trehalose utilization variants on CDI outcome we combined the three variants into a single predictor of severe CDI outcome and performed a logistic regression conditioned on severe outcome risk strata. When combined, the presence of any of the three trehalose utilization variants does not significantly affect the odds of severe CDI outcome (OR, 1.10; 95%CI, 0.61-1.99; *P*=0.86). To evaluate whether these results translated to the original unmatched cohort, we also tested for associations between trehalose utilization variants and severe infections using severe risk score as a covariate rather than matching criterion. To determine trehalose utilization genotypes in the larger cohort for which genomic data was unavailable, we performed an imputation where the presence of trehalose utilization variants (TreR C171S, TreR L172I, and the four gene trehalose insertion) was assumed to have the same distribution across ribotypes as was observed in the matched cohort (see Supplementary Methods). We performed a logistic regression modeling severe outcome with presence of any of the three trehalose utilization variants as a predictor and severe risk score as a covariate. When combined, the presence of any of the three trehalose utilization variants does not significantly affect the odds of severe CDI outcome (Median OR, 0.99; Range, 0.87-0.99; Median *P*=0.97; Range, 0.64-0.99; N=586).

## DISCUSSION

The ability to predict the clinical outcome of *C. difficile* infection based on patient and pathogen characteristics could help guide therapy for this important nosocomial infection. An enhanced ability to utilize trehalose was shown to be associated with increased virulence in mice, prompting us to evaluate this relationship using a case-control study performed with a large clinical data set. Our controlled analysis failed to detect a significant association between any of the previously described trehalose utilization variants and severe CDI outcome. Our results from patients infected with a diversity of ribotypes are consistent with Eyre et al.’s observed lack of association between the four gene trehalose insertion and 30-day all-cause mortality specifically in patients infected with RT015. We also observed that the four-gene trehalose insertion is not exclusive to RT078, but rather is found throughout the *C. difficile* phylogeny.

The lack of association between trehalose utilization variants and severe CDI outcome in hospitalized patients emphasizes the need to incorporate clinical results earlier into the genetic variant discovery pipeline. Indeed, a more relevant hypothesis generating process could begin by identifying genetic loci of interest first through comparative genomic analysis of clinical isolates followed by validation in lab strains with molecular and animal studies. A critical component of this discovery work is to control for both patient factors and strain background as both may confound analysis, which we did above in separate analyses using two distinct modeling strategies. Given the association between ribotype, recent acquisition, and CDI outcome, controlling for patient factors is critical for identifying the genetic variants associated with severe CDI outcome[12].

While these clinical data cannot rule out a potential role of trehalose utilization variants in the success of hypervirulent *C. difficile* lineages, they do emphasize the difference between human infection and murine models. More broadly, generating hypotheses in controlled laboratory systems and having them generalize to genetically and clinically heterogeneous human populations will limit insights to only the most penetrant phenotypes. With increasing availability of electronic health record and microbial genomic data, it is now becoming feasible to flip the script and generate hypotheses through analysis of human data, which can subsequently be tested in appropriate animal and *in vitro* systems with *a priori* knowledge of relevance to human disease.

## Data Availability

Data are available both in the supplement and in the github repository: https://github.com/katiesaund/clinical_cdifficile_trehalose_variants

## ACKNOWLEDGEMENTS

We thank Ali Pirani for bioinformatics support. We thank Arianna Miles-Jay for her helpful suggestions.

## REFERENCES

1. Clifford McDonald L, Coignard B, Dubberke E, Song X, Horan T, Kutty PK. Recommendations for Surveillance of Clostridium difficile–Associated Disease. Infect Control Hosp Epidemiol 2007; 28.

2. Rao K, Micic D, Natarajan M, et al. Clostridium difficile Ribotype 027: Relationship to Age, Detectability of Toxins A or B in Stool with Rapid Testing, Severe Infection, and Mortality. Clin Infect Dis 2015; 61:233–241. Available at: http://www.ncbi.nlm.nih.gov/pubmed/25828993. Accessed 10 November 2016.

3. Walker AS, Eyre DW, Wyllie DH, et al. Relationship between bacterial strain type, host biomarkers, and mortality in clostridium difficile infection. Clin. Infect. Dis. 2013; 56:1589–1600. Available at: https://oup.silverchair-cdn.com/oup/backfile/Content_public/Journal/cid/56/11/10.1093_cid_cit127/2/cit127.pdf?Expires=1500592769&Signature=btVMYEpwYmM7E08K3gthf-e6qsNyxMnPaDF8UHR3eTBis0v4jD0zfPQobzNDM7c604DF7gHqUQv-HzZswhVkCQ1HmGjiMR01OWGQT~M~4Tcy1XBCWIXs. Accessed 19 July 2017.

4. Collins J, Robinson C, Danhof H, et al. Dietary trehalose enhances virulence of epidemic Clostridium difficile. Nature 2018; 553:291–294. Available at: https://www.nature.com/articles/nature25178.pdf. Accessed 12 April 2019.

5. Eyre DW, Didelot X, Buckley AM, et al. Clostridium difficile trehalose metabolism variants are common and not associated with adverse patient outcomes when variably present in the same lineage. EBioMedicine 2019; Available at: https://linkinghub.elsevier.com/retrieve/pii/S2352396419302774.

6. R Core Team. R: A language and environment for statistical computing. 2018; Available at: https://www.r-project.org/.

7. Rao K, Micic D, Natarajan M, et al. Clostridium difficile Ribotype 027: Relationship to Age, Detectability of Toxins A or B in Stool with Rapid Testing, Severe Infection, and Mortality. Clin Infect Dis 2015; 61:233–241.

8. Andrews Si. FastQC: a quality control tool for high throughput sequence data. 2010;

9. Bolger AM, Lohse M, Usadel B. Trimmomatic: A flexible trimmer for Illumina sequence data. Bioinformatics 2014; 30:2114–2120. Available at: http://www.usadellab.org/cms/index.php?page¼trimmomatic. Accessed 1 March 2018.

10. Page AJ, Cummins CA, Hunt M, et al. Roary: Rapid large-scale prokaryote pan genome analysis. Bioinformatics 2015; 31:3691–3693.

11. Collins J, Danhof H, Britton RA. The role of trehalose in the global spread of epidemic Clostridium difficile. Gut Microbes 2018; :1–6. Available at: http://www.tandfonline.com/action/journalInformation?journalCode=kgmi20. Accessed 21 August 2018.

12. Martin JSH, Eyre DW, Fawley WN, et al. Patient and strain characteristics associated with clostridium difficile transmission and adverse outcomes. Clin Infect Dis 2018; 67:1379–1387. Available at: https://academic.oup.com/cid/article-abstract/67/9/1379/4969186. Accessed 1 May 2019.

